# A Mini Review on Current Clinical and Research Findings for Children Suffering from COVID-19

**DOI:** 10.1101/2020.03.30.20044545

**Authors:** Xiao Li, Kun Qian, Ling-ling Xie, Xiu-juan Li, Min Cheng, Li Jiang, Björn W. Schuller

**Author notes:** Corresponding author: Dr. Li Jiang.

## Abstract

**Background:** As the novel coronavirus triggering COVID-19 has broken out in Wuhan, China and spread rapidly worldwide, it threatens the lives of thousands of people and poses a global threat on the economies of the entire world. However, infection with COVID-19 is currently rare in children.

**Objective:** To discuss the latest findings and research focus on the basis of characteristics of children confirmed with COVID-19, and provide an insight into the future treatment and research direction.

**Methods:** We searched the terms “COVID-19 OR coronavirus OR SARS-CoV-2” AND “Pediatric OR children” on PubMed, Embase, Cochrane library, NIH, CDC, and CNKI. The authors also reviewed the guidelines published on Chinese CDC and Chinese NHC.

**Results:** We included 25 published literature references related to the epidemiology, clinical manifestation, accessary examination, treatment, and prognosis of pediatric patients with COVID-19.

**Conclusion:** The numbers of children with COVID-19 pneumonia infection are small, and most of them come from family aggregation. Symptoms are mainly mild or even asymptomatic, which allow children to be a risk factor for transmission. Thus, strict epidemiological history screening is needed for early diagnosis and segregation. This holds especially for infants, who are more susceptible to infection than other age groups in pediatric age, but have most likely subtle and unspecific symptoms. They need to be paid more attention to. CT examination is a necessity for screening the suspected cases, because most of the pediatric patients are mild cases, and plain chest X-ray do not usually show the lesions or the detailed features. Therefore, early chest CT examination combined with pathogenic detection is a recommended clinical diagnosis scheme in children. The risk factors which may suggest severe or critical progress for children are: Fast respiratory rate and/or; lethargy and drowsiness mental state and/or; lactate progressively increasing and/or; imaging showed bilateral or multi lobed infiltration, pleural effusion or rapidly expending of lesions in a short period of time and/or; less than 3 months old or those who underly diseases. For those critical pediatric patients with positive SARS-CoV-2 diagnosis, polypnea may be the most common symptom. For treatment, the elevated PCT seen in children in contrast to adults suggests that the underlying coinfection/secondary infection may be more common in pediatric patients and appropriate antibacterial treatment should be considered. Once cytokine storm is found in these patients, anti-autoimmune or blood-purifying therapy should be given in time. Furthermore, effective isolation measures and appropriate psychological comfort need to be provided timely.

## Introduction

Since late December 2019, the numbers of unknown registered cases of pneumonia originating from a novel coronavirus had outbroken in Wuhan, Hubei province, China. A fast outbreak could be observed rapidly reaching other provinces and cities of China, and subsequently, to many countries around the three big oceans. Now, the epidemic of the 2019 novel type of coronavirus (SARS-CoV-2) has threatened the lives of thousands of people and posed a global threat on the economics of the whole world. On 11 February 2020, the World Health Organization (WHO) named the disease related to SARS-CoV-2 officially as the 2019 coronavirus disease (COVID-19) ^[1-3].^

Coronaviruses (CoV) are members of the subfamily Coronavirinae in the Coronaviridae family, Nidovirales order. CoV consists of four genera: α, β, γ, and δ coronaviruses. The alpha- and beta-coronaviruses infect only mammals, while the gamma- and delta-coronaviruses mainly infect birds. COVID-19 belongs to the β genus coronaviruses ^[1]^. SARS-CoV-2 is a novel human coronavirus (HCoVs) besides four HCoVs ((α-coronaviruses (229E and NL63), β-coronaviruses (OC43 and HKU1)), Middle East respiratory syndrome coronavirus (MERSr-CoV), as well as the severe acute respiratory syndrome-related coronavirus (SARS-CoV) ^[1]^. It is an enveloped positive single-stranded RNA virus with 60–140 nm in diameter. The shape of SARS-CoV-2 is spherical or elliptical ^[4,5]^. The consistency of whole genome sequence of SARS-CoV-2 with SARS-like coronavirus in bats (bat-SL-CoVZC45) ranges from 86.9% ^[5]^ to 89% ^[6]^. “The SARS-CoV-2 is sensitive to ultraviolet light and heat, and it can be inactivated at 56°C for 30 minutes. Lipid solvents, except chlorahexidine, such as ethyl ether, and 75 % ethanol can also effectively inactivate the virus” ^[2,4]^.

Up until 22 March 2020, there have been 333,751 cases confirmed with COVID-19 and 14,524 people died of it worldwide (data from the WHO). However, cases of pediatric patients infected with COVID-19 are currently rare – even though the situation of COVID-19 has become more urgent and severe around the world. Thus far, literature on children with COVID-19 is comparably rare.

To help better understand how it would affect children and what is the latest specific clinical and research finding on children with it, we provide a mini-review based on 25 literature references covering the fields of epidemiology, clinical manifestation, accessary examination, treatment, and prognosis of pediatric patients with COVID-19. The selection process is described in detail in the next section.

### Method

We have searched the query string “COVID-19 OR coronavirus OR SARS-CoV-2” AND “Pediatric OR children” (publication date was restricted in 1 year) on PubMed, Embase, Cochrane library, NIH, CDC, and CNKI by 16 March 2020. We yielded 85 articles on PubMed, 1 on Embase, none on Cochrane library, 8 on CNKI, none on Cochrane library, NIH, and CDC. Subsequently, we went over the abstracts of those articles to see whether they were related to the current pandemic and ruled out overlapping literature. Ultimately, we ended up with 25 articles (17 in English, 8 in Chinese – cf. references) including 3 reviews to read the full article. Afterwards, we excluded those with relatively lower confidence level compared to literature on the same topic. In addition, we reviewed the guidelines related to COVID-19 published on the Chinese CDC and Chinese NHC. Note that at the time, literature on the topic is growing rapidly, and search results may differ quickly. In the following, we provide the summary obtained.

## Epidemiological features

### Transmission portal

According to these published retrospective studies and guidelines, “the main source of infection are patients infected with COVID-19 with or without clinical symptoms” ^[4]^. Furthermore, during the period of incubation, those infected also have the ability to transmit the virus ^[6,14]^.

Normally, COVID-19 is spread through respiratory droplets. Close contact with symptomatic or asymptomatic patients with a positive COVID-19 test is also a route of transmission. Moreover, current research comes to the conclusion that SARS-CoV-2 can be transmitted via the fecal-oral pathway as well ^[4]^. However, mother–infant vertically or breast milk transmission remains unclear. It is reported that the youngest individual with a COVID-19 diagnosis is only 36 hours old. In addition, two neonates were diagnosed with COVID-19. Thus, vertical-transmission appears possible ^[11]^.

### Susceptible population

Normally, humans are generally susceptible to SARS-CoV-2. Current obersvation has it that, among the patients confirmed with SARS-CoV-2 (44,653 cases) until February 11, 2020, in China, 2.1% were between 0-19 years old, which suggested that the population of pediatric patients were rather small ^[9]^. Moreover, the epidemiological data showed that children who suffered from a SARS-CoV-2 infection were mainly resulting from familial aggregation ^[4,9]^. The epidemiological study of HCoV among children based on 3,883 pharyngeal swab samples in 2015 also showed that the positive rate of HCoV decreased with age, and the positive rate of HCV DNA was highest in the < 1 year old group ^[17]^.

According to the data of the Dong Y’ group, which focused on 2,143 pediatric patients with positive SARS-CoV-2 tests forwarded to the Chinese Center for Disease Control and Prevention, young individuals of any age were susceptible to SARS-CoV-2. However, younger children, particularly infants, were more at risk of SARS-CoV-2 infection ^[10]^, while neonates – especially preterm infants – show mostly insidious and non-specific symptoms ^[11]^.

## Clinical characteristics

Like many other pediatric respiratory systems infectious disease, at the onset of COVID-19, not specifically, children affected by SARS-CoV-2 mainly present asymptomatic or only mild symptoms of the respiratory or gastrointestinal systems such as fever, cough, mild diarrhea, or headache. The majority of the young patients had low to medium fever, or even none ^[4,9]^.

Dyspnea, shortness of breath, nostrils flaring, cyanosis, moist rales on auscultation, and other pneumonia symptoms may appear with the progress of COVID-19 alongside toxic symptoms of systemic infection, such as lethargy, mental fatigue, reduced appetite, and others. Critical patients may manifest a respiratory failure largely invariant to oxygen treatment; moreover, even septic shock, multiple organ failure, and coagulation dysfunction may occur ^[3,4,8,18]^.

According to the current literature on the pediatric cases, children confirmed with COVID-19 mostly had good prognosis, with considerably less severe to critical progress (5.9%) as compared to adult patients (18.5%). This suggests that, compared with adult patients, clinical manifestations of pediatric patients with positive SARS-CoV-2 tests may be milder ^[4,10]^.

### Accessory examinations

#### Laboratory testing

According to the diagnosis and treatment recommendations by the National Center for children’s health and disease, “routine blood test white blood cell count of children with SARS-CoV-2 is usually normal or decreased, with generally lower lymphocyte count; severe cases may exhibit progressive lymphocytopenia. C-reactive protein (CRP) is mostly normal or increased. Procalcitonin (PCT) is also at normal level in most cases. Elevation of liver enzymes, muscle enzymes and myoglobin, and increased level of D-dimer could be seen in severe cases” ^[4]^.

#### Nucleic acid testing

is the gold standard of laboratory diagnosis. SARS-CoV-2 nucleic acid can be detected by RT-PCR or by viral gene sequencing of upper airway specimens (pharyngeal/nasal swabs) and lower airway specimens (sputum, bronchoalveolar lavage fluid), urine, stool, conjunctival secretions, or blood samples. Other methods include that isolation of SARS-CoV-2 particles from human cells by viral cultures ^[4]^.

#### Imaging features

*“* Given chest X-ray examination in the early stage of pneumonia cases, chest images show multiple small patchy shadows and interstitial changes, remarkable in the lung periphery. In severe cases, bilateral multiple ground-glass shadows, infiltrative opacity and pulmonary consolidation may develop, accompanied by rare pleural effusion” ^[4]^.

However, because most pediatric patients with COVID-19 belong to the mild category, the plain chest X-ray of them usually do not capture the lesions detailed characteristics of the lungs. In the result, diagnoses may be taken wrongly. To circumvent this, chest CT seems crucial to be taken early on ^[8]^.

In fact, chest CT scanning can well show pulmonary lesions due to the high sensitivity and tomography. In bilateral lungs, it provides segmental consolidation as well as ground glass opacity (GGO). In those young patients who are severely infected, several lobar can be affected ^[4]^.

Additionally, for infants, especially preterm babies, abdominal radiography may exhibit the characteristic intestinal ileus ^[11]^.

### Diagnostic criteria

COVID-19 infection should be taken as possibility once a criterium is met within the epidemiological history and two of the clinical manifestations in ^[4]^ hold.

A positive diagnosis can be stated if: “1) Throat swab, sputum, stool, or blood samples tested positive for SARS-CoV-2 nucleic acid using RT-PCR; 2) genetic sequencing of throat swab, sputum, stool, or blood samples are being highly homologous with the known SARS-CoV-2; 3) SARS-CoV-2 granules are being isolated by viral culture from throat swab, sputum, stool, or blood specimens^” [4]^.

For those suspected patients with negative nucleic acid detection, for identification of serum antibody holds: positive serum SARS-CoV-2 -specific IgM, or specific IgG antibody titer during recovery are at minimum four times above the level in the acute phase ^[2,4]^.

### Clinical classifications

COVID-19 was categorized by clinical manifestations, lab test outcome, and chest X-ray/CT imaging into five categories: firstly, asymptomatic infection, followed by mild, and moderate, reaching to severe, and critical.

The diagnostic criteria are referred to in ^[2,4]^.

### Management principles

In terms of management principles, one can stress the importance of “early identification”, “early isolation”, “early diagnosis”, and “early treatment” ^[4]^.

Generally, without certain effective cure, treatments of COVID-19 consist of both symptomatic and respiratory supports. For now, antiviral treatments (virazole, oseltamivir, and interferon) are the standardised drugs for COVID-19 patients. Application of countermeasures reported so far includes usage of antibiotics, immunoglobulin, intravenous steroid, and TCM.

***In mild cases, one should refrain from applying*** broad-spectrum antibiotics or corticosteroids” ^[4]^.

***Further, in severe as well as critical cases***, *“* Antibiotics, corticosteroids, bronchoalveolar lavage, mechanical ventilation, and other more invasive intervention, such as blood purification and extracorporeal membrane oxygenation (EMCO) should be applied cautiously, based on cost-benefit evaluation” ^[4]^.

***Finally, as to discharge, the young patients need to show a return to their normal temperature of the body for a minimum of three consecutive days***, significant recovery of their respiratory system, and succeed in having two consecutive, but at least one full day apart negative test results for respiratory pathogenic nucleic acid. Depending on the situation, home isolation is subsequently advised over two full weeks. ^[4,11]^.

### Prognosis

Most asymptomatic or mild patients had a good prognosis. So far, there is one registered death case of a child patient. In addition, child COVID-19 cases were mild, with a one to two week recovery time subsequent to onset ^[9]^. However, severe and critical cases may be left with long-term lung damage ^[19].^ Brain damage such as chronic demyelination and seizures as comorbidity of pediatric patients with HCoV have also been reported ^[15]^.

## Discussion

To our best knowledge on COVID-19, among the patients confirmed with COVID-19, the population of pediatric patients is rather small ^[4]^. The children confirmed with COVID-19 mostly have good prognosis. A single child patient passed away; for other child patients, progression of the disease was observed as mild ^[9,10]^. Yet, in more general, infectious diseases often tend to behave less aggressively in child patients. For severe respiratory distress syndrome (SARS), the total death ratio is reported within 7% to 17%. For individuals with less than 24 years of age, the death ratio is 0% compared to 50% mortality rates in adults older than 65 years ^[9]^. Looking at those cases of COVID-19 judged as severe, children’s rate is also considerably below adults’ rate (49.0%, 1023/2087) ^[19]^. The main explanation would be:1. One possible reason is that children have been protected well resulting in lower potential exposure with the virus due to considerably lower travelling track record and potentially relatively higher indoor time. 2. Children possess more active innate immune systems, including mostly fully functional lungs owing to low exposure to smoking induced and other air pollution. 3. Children do not really have complex underlying disorders. 4.The differences in distribution, maturation and function of viral receptor angiotensin converting enzyme II (ACE2) are usually named a potential cause for age-based differences in infection rate. ACE2 was already considered a cell receptor for SARS-CoV^[9]^. Recent reports indicate that ACE2 in ciliated bronchial epithelial cells and type II pneumocytes is also likely the binding receptor for SARS-CoV-2. A usual theory for children’s lower sensitivity to SARS-CoV-2 is based on their less maturity and lower function of ACE2 compared with that in adults ^[9]^.

Nevertheless, the children, especially infants, who are confirmed with COVID-19 could be an insidious infectious source because of their asymptomatic or mild symptomatic infection due to a weak immune system or their own physical characteristics. A largely asymptomatic infant with COVID-19 with high viral load was reported in Singapore, and this case highlights the difficulties in establishing the true incidence of COVID-19, as asymptomatic individuals may play important roles in human-to-human transmission in the community ^[13]^. A study focused on 24 asymptomatic infections identified that the infection period of asymptomatic patients may be as long as three weeks, and the infected patients may develop into serious diseases ^[14]^. Furthermore, the bad habits of young children such as sucking hands, putting their hands all around and unwilling to wash hands could be important risk factors in contact transmission for children. Thus, it is necessary to guarantee family daily prevention to keep children from COVID-19 infection and assure awareness of the importance of its early diagnosis. Also, during the epidemic period, children’s health clinics and vaccination clinics should be reduced reasonably, and parent-child activities, kindergartens, primary schools, and other group activities related to children should be suspended. In clinical case, screening was mostly based on the epidemiological track record. Additional basis were raised body temperature and symptoms of the respiratory tract. For children, it is more important to trace close contact history, and pathogen examination needs to be timely executed along with longitudinal monitoring through viral nucleic acid detection. Further isolation recommendations and ongoing nucleic acid testing are also recommended for discharged patients ^[14]^. Additionally, it appears crucial to follow up with infants’respiratory as well as gastrointestinal symptoms and vital indications steadily, which is the most susceptible population in children, at mild onset that could progress into severe at a later stage ^[11]^.

Moreover, for screening the suspected cases, since “most of the pediatric patients are mild cases, plain chest X-ray often fails to show the lesions or the detailed features, leading to misdiagnosis or missed diagnosis” ^[4,8,16]^. As a consequence, chest CT early on is seen mandatory.

Also, a number of suspected pediatric patients with a negative nucleic acid of SARS-CoV-2 virus were rightfuly considered potential infection cases in accordance with usual chest CT visible lesions. This further encourages early diagnosis and treatment, but also isolating of infected children in effective ways ^[8]^.

In a recent report, the authors studied the CT features of 20 children, and came to the conclusion that consolidation with surrounding halo signs are frequent in children with COVID-19. Interestingly, these are dissimilar from those of grown-ups. One may assume such consolidation with surrounding halo signs as a distinctive feature of young individuals affected ^[8]^. Another study reported the CT results of 3 children with positive COVID-19 diagnosis which showed patchy ground-glass opacities that were similar, but more modest compared to reports in adults ^[18]^.However, the samples of both studies are indeed small, in the future, large-scale studies focusing on the imaging features of children with COVID-19 are needed.

Furthermore, the data from the first group also showed that the PCT has significantly increased in 80% of the cases in this study with or without coinfection evidence existing, which is not common in adult patients ^[8]^. This suggests that the underlying coinfection or secondary infection may be more common in pediatric patients. If a secondary infection/coinfection is suspected, a collection of the specimen should be executed to detect potential secondary bacterial and/or fungal infections, and routine antibacterial treatment also should be considered in this situation ^[2,8]^.

In 2017, a large-scale retrospective research focusing on epidemiology as well as on clinical attributes of HCoVs (HKU1, 229E, OC43, and NL63) in the pediatric population (261 cases) showed the risks linked to aggravation of infection. The risk factors were: younger than two years of age and in addition cardiovascular, genetic or congenital and respiratory chronic complex conditions which in the context of requiring respiratory support. Genetic or congenital underlying disorders were associated with PICU admission ^[17]^. For those critical pediatric patients with positive COVID-19 diagnosis, the most frequently observed symptoms were polypnea, then fever, and finally coughing ^[19]^.

Some severe or critical children were found to have an excessive active immune response, which may result in long-term lung injury as well as fatal health problems. Coronaviruses including SARS-CoV-2, SARS, and MERS were proved to have the potential ability to lead to significant and peculiar destructive host immune response that is related to severe lung damage. Researches exhibited the lung impairment is mostly observed in patients in a critical state, linked to a cytokine storm, marked by raised levels of pro-inflammatory cytokines (IL-1β, IL-6, IL-12, TNF, IFN-γ), as well as of anti-inflammatory cytokines (IL-4, IL-10, IL-13, TGF-β) ^[12,19-25]^.

These findings suggest that it is necessary to detect the indicator of the inflammatory and immune status, which could help doctors to assess the clinical progress, be on alert for severe and critical cases, and provide anti-autoimmune or blood-purifying therapy in time, once cytokine storm occurs ^[2,19]^.

Early identification and diagnosis of the severe/critical pediatric patients are always the priority to result in appropriate treatment and good prognosis.

In the latest version of *“Scheme for Diagnosis and Treatment of the 2019 Novel Coronavirus Pneumonia”*, pediatric patients with “fast respiratory rate and/or; lethargy and drowsiness mental state and/or; lactate progressively increasing and/or; imaging showed bilateral or multi lobed infiltration, pleural effusion or rapidly expending of lesions in a short period of time and/or; less than 3 months old or those which underly diseases (congenital heart disease, bronchopulmonary dysplasia, respiratory tract deformity, abnormal hemoglobin, severe malnutrition, etc.) and/or, immune deficiency/hypofunction (long-term use of immunosuppressant drugs)”, are in high risk progressing into severe to critical illness, and intravenous immunoglobulin may be considered in those children ^[2]^.

For those children isolated alone from their parents, maternal separation may cause anxiety and depression to infants or children. For this reason, their psychological care-taking for their wellbeing needs to be assured. All of the treatment plan and segregation situation have to be discussed with the responsible care-takers of the young individuals. Psychologists or suited related experts are required to help cease their anxiety and panic.

In conclusion, the numbers of children with COVID-19 pneumonia infection are small, and most of them come from family aggregation. Symptoms are mainly mild or even asymptomatic which allow children to be a risk factor for transmission. Thus, strict epidemiological history screening is needed for early diagnosis and segregation. This holds especially for infants, who are more susceptible to infection than other age groups in pediatric age, but have most likely subtle and unspecific symptoms. They need to be paid more attention to. CT examination is a necessity for screening the suspected cases, because the majority of the young COVID-19 infected are classified as mild infection, and plain chest X-ray mostly gives not sufficient insight into the lesions or lung details. Therefore, early chest CT examination combined with pathogenic recognition is a recommended clinical diagnosis scheme in children. The risk factors which may suggest severe or critical progress for children are: Fast respiratory rate and/or; lethargy and drowsiness mental state and/or; lactate progressively increasing and/or; imaging showed bi- or multi-lobed infiltration, pleural effusion, or rapidly expending of lesions in a short period of time and/or; less than 3 months old or those who underly diseases. For those critical pediatric patients with positive SARS-CoV-2 diagnosis, polypnea may be the most common symptom. For treatment, the elevated PCT seen in children in contrast to adults suggests that the underlying coinfection/secondary infection may be more common in pediatric patients and appropriate antibacterial treatment should be considered. Once cytokine storm is found in these patients, anti-autoimmune or blood-purifying therapy should be given in time. Finally, it is of crucial importance to assure early isolation combined with best possible psychological support for the young patients.

## Data Availability

Does not apply.

## Acknowledgement

This work was partially supported by the Zhejiang Lab’s International Talent Fund for Young Professionals (Project HANAMI), P. R. China, the JSPS Postdoctoral Fellowship for Research in Japan (ID No. P19081) from the Japan Society for the Promotion of Science (JSPS), Japan, the Grants-in-Aid for Scientific Research (No. 19F19081) from the Ministry of Education, Culture, Sports, Science and Technology (MEXT), Japan, and the EU’s HORIZON 2020 Grant No.115902 (RADAR CNS).

## Reference

[1] Cui J, Li F, Shi ZL. Origin and Evolution of Pathogenic Coronaviruses. Nat Rev Microbiol. 2019 Mar;17(3):181–192. doi: 10.1038/s41579-018-0118-9.

[2] National Health Commission of the People’s Republic of China. Scheme for Diagnosis and Treatment of 2019 Novel Coronavirus Pneumonia (The 7th Trial Edition), (2020-03-03, e-pub online).

[3] Department of Maternal and Child Health of the National Health Commission of the People’s Republic of China. Scheme for novel coronavirus infection prevention and control for children and pregnant women (2020-02-02, e-pub online)

[4] Chen ZM, Fu JF, Shu Q, Chen YH, Hua CZ, Li FB, Lin R, Tang LF, Wang T L, Wang W, Wang YS, Xu WZ, Yang ZH, Ye S, Yuan TM, Zhang CM, & Zhang YY. Diagnosis and treatment recommendations for pediatric respiratory infection caused by the 2019 novel coronavirus. World J Pediatr. 2020 Feb 5. [Online ahead of print] doi: 10.1007/s12519-020-00345-5.

[5] Zhu N, Zhang D, Wang W, Li X, Yang B, Song J, Zhao X, Huang B, Shi W, Lu R, Niu P, Zhan F, Ma X, Wang D, Xu W, Wu G, Gao GF, Tan W, & China Novel Coronavirus Investigating and Research Team. A Novel Coronavirus from Patients with Pneumonia in China, 2019. N Engl J Med. 2020,382:727–733. doi: 10.1056/NEJMoa2001017.

[6] Chan JF, Yuan S, Kok KH, To KK, Chu H, Yang J, Xing F, Liu J, Yip CC, Poon RW, Tsoi HW, Lo SK, Chan KH, Poon VK, Chan WM, Ip JD, Cai JP, Cheng VC, Chen H, Hui CK, Yuen KY. A familial cluster of pneumonia associated with the 2019 novel coronavirus indicating person-to-person transmission: a study of a family cluster. Lancet. 2020,395:514–523. doi: 10.1016/S0140-6736(20)30154-9.

[7] Kong W, Wang Y, Zhu H, Lin X, Yu B, Hu Q, Guo D, Peng J. Epidemiological characteristics of human coronaviruses among children in Wuhan, 2008-2013. Zhonghua Yu Fang Yi Xue Za Zhi. 2015,49:444–6.

[8] Xia W, Shao J, Guo Y, Peng X, Li Z, Hu D. Clinical and CT features in pediatric patients with COVID-19 infection: Different points from adults. Pediatr Pulmonol. 2020,1–6. doi: 10.1002/ppul.24718.

[9] Lee PI, Hu YL, Chen PY, Huang YC, Hsueh PR. Are children less susceptible to COVID-19? J Microbiol Immunol Infect. 2020 Feb 25. [Online ahead of print]. doi: 10.1016/j.jmii.2020.02.011.

[10] Dong Y, Mo X, Hu Y, Qi X, Jiang F, Jiang Z, Tong S. Epidemiological characteristics of 2143 pediatric patients with 2019 coronavirus disease in China. Pediatrics. 2020 Mar 16. [Online ahead of print]. doi: 10.1542/peds.2020-0702

[11] Wang L, Shi Y, Xiao T, Fu J, Feng X, Mu D, Feng Q, Hei M, Hu X, Li Z, Lu G, Tang Z, Wang Y, Wang C, Xia S, Xu J, Yang Y, Yang J, Zeng M, Zheng J, Zhou W, Zhou X, Zhou X, Du L, Lee SK, Zhou W; Working Committee on Perinatal and Neonatal Management for the Prevention and Control of the 2019 Novel Coronavirus Infection. Chinese expert consensus on the perinatal and neonatal management for the prevention and control of the 2019 novel coronavirus infection (First edition). Ann Transl Med. 2020,8: 47.doi: 10.21037/atm.2020.02.20.Review.

[12] Han Q, Lin Q, Jin S, You L. Coronavirus SARS-CoV-2: A brief perspective from the front line. J Infect. 2020 Feb 25. [Online ahead of print]. doi: 10.1016/j.jinf.2020.02.010. Online ahead of print.

[13] Kam KQ, Yung CF, Cui L, Lin Tzer Pin R, Mak TM, Maiwald M, Li J, Chong CY, Nadua K, Tan NWH, Thoon KC. A Well Infant with Coronavirus Disease 2019 (COVID-19) with High Viral Load. Clin Infect Dis. 2020 Feb 28. [Online ahead of print]. doi: 10.1093/cid/ciaa201.

[14] Hu Z, Song C, Xu C, Jin G, Chen Y, Xu X, Ma H, Chen W, Lin Y, Zheng Y, Wang J, Hu Z, Yi Y, Shen H. Clinical characteristics of 24 asymptomatic infections with COVID-19 screened among close contacts in Nanjing, China Sci China Life Sci. 2020 Mar 4. [Online ahead of print]. doi: 10.1007/s11427-020-1661-4.

[15] Jacomy H, Fragoso G, Almazan G, Mushynski WE, Talbot PJ. Human coronavirus OC43 infection induces chronic encephalitis leading to disabilities in BALB/C mice. Virology. 2006,349:335–46. DOI: 10.1016/j.virol.2006.01.049

[16] Li W, Cui H, Li K, Fang Y, Li S. Chest computed tomography in children with COVID-19 respiratory infection. Pediatr Radiol. 2020 Mar 11. [Online ahead of print]. doi: 10.1007/s00247-020-04656-7.

[17] Varghese L, Zachariah P, Vargas C, LaRussa P, Demmer RT, Furuya YE, Whittier S, Reed C, Stockwell MS, Saiman L. Epidemiology and Clinical Features of Human Coronaviruses in the Pediatric Population. J Pediatric Infect Dis Soc. 2018,7:151–158. doi: 10.1093/jpids/pix027.

[18] Ji LN, Chao S, Wang YJ, Li XJ, Mu XD, Lin MG, Jiang RM. Clinical features of pediatric patients with COVID-19: a report of two family cluster cases. World J Pediatr. 2020 Mar 16. [Online ahead of print]. doi: 10.1007/s12519-020-00356-2.

[19] Sun D, Li H, Lu XX, Xiao H, Ren J, Zhang FR, Liu ZS. Clinical features of severe pediatric patients with coronavirus disease 2019 in Wuhan: a single center’s observational study. World J Pediatr. 2020 Mar 19. [Online ahead of print]. doi: 10.1007/s12519-020-00354-4.

[20] Wu Z, McGoogan JM. Characteristics of and important lessons from the coronavirus disease 2019 (COVID-19) outbreak in China: summary of a report of 72,314 cases from the Chinese Center for Disease Control and Prevention. JAMA. 2020 Feb 24. [Online ahead of print].https://doi.org/10.1001/jama.2020.2648

[21] Huang C, Wang Y, Li X, Ren L, Zhao J, Hu Y, Zhang L, Fan G, Xu J, Gu X, Cheng Z, Yu T, Xia J, Wei Y, Wu W, Xie X, Yin W, Li H, Liu M, Xiao Y, Gao H, Guo L, Xie J, Wang G, Jiang R, Gao Z, Jin Q, Wang J, Cao B. Clinical features of patients infected with 2019 novel coronavirus in Wuhan, China. Lancet. 2020,15;395(10223):497–506. doi: 10.1016/S0140-6736(20)30183-5.

[22] Zumla A, Hui DS, Azhar EI, Memish ZA, Maeurer M. Reducing mortality from 2019-nCoV: host-directed therapies should be an option. Lancet. 2020,22;395(10224):e35–e36. doi: 10.1016/S0140-6736(20)30305-6.

[23] Hui DSC, Zumla A. Severe acute respiratory syndrome: historical, epidemiologic, and clinical features. Infect Dis Clin N Am. 2019, 33:869–89. doi: 10.1016/S0140-6736(20)30183-5.

[24] Zumla A, Hui DS, Perlman S. Middle East respiratory syndrome. Lancet. 2015,5;386(9997):995–1007. doi: 10.1016/S0140-6736(15)60454-8.

[25] Li G, Fan Y, Lai Y, Han T, Li Z, Zhou P, Pan P, Wang W, Hu D, Liu X, Zhang Q, Wu J. Coronavirus infections and immune responses. J Med Virol. 2020 Apr;92(4):424–432. doi: 10.1002/jmv.25685.

